# Factors on Knowledge, Health Beliefs, and Willingness toward Human Papillomavirus Vaccine among Chinese Parents, a cross-sectional study in Ordos City, China

**DOI:** 10.1101/2025.08.28.25334402

**Authors:** Sensen Tan, Yufei Li, Sumeng Wang, Huijiao Yan, Yang Xu, Linlin Zhang, Jiao Xue, Xuemei Lian, Jian Yin, Youlin Qiao

## Abstract

**Background:** This cross-sectional study was conducted in Ordos City aimed to explore the potential factors influencing parents’ awareness, knowledge, and health beliefs toward human papillomavirus (HPV), and willingness to vaccinate their 13-18-year-old daughters against HPV free of charge.

**Methods:** This cross-sectional study was conducted on parents with daughters aged 13-18 years, in Ordos City, China. Data was collected using an online questionnaire survey, including parental sociodemographic characteristics, awareness, knowledge, and health beliefs about HPV, and willingness to HPV vaccine. Mean scores on HPV and its vaccine-related knowledge and were calculated, separately, and parents above the mean level were included in bivariate and multivariate analyses.

**Results:** After removing illogical values, a total of 1,547 parents were analyzed, with an average age of 42.25 years old, and most of them (92.4%) were female. 78% of participants had heard of HPV, 93% had heard of the HPV vaccine, and 85.5% of parents were willing to vaccinate their daughters. The mean Knowledge score (KS) was 1.98 ± 1.81 (out of 7) and the mean Health belief score (HBS) was 6.18 ± 2.94 (out of 12). Factors that potentially influence knowledge were found to be gender, married status, education, and income. Registered permanent residence (girls), income, and vaccination status (female parents) were significantly associated with health beliefs and willingness.

**Conclusions:** In conclusion, many parents had insufficient knowledge about the vaccine and low health beliefs. It is critical to conduct health education campaigns to abolish the barriers identified to accelerate the rollout and increase the national vaccination coverage of the HPV vaccine in China.

**Synopsis:** A cross-sectional study in Ordos City, China, reveals parents’ limited knowledge and low health beliefs regarding HPV vaccines, highlighting the need for effective health education.

## 1 Introduction

Cervical cancer is one of the most common malignant tumors in women. In 2020, there were 604,127 new cases and 341,831 deaths of cervical cancer worldwide, among which, approximately 110,000 new cases and 59,000 deaths were from China^1^. Cervical cancer is caused by persistent infection with high-risk human papillomavirus (HPV)^2^,which is one of the most prevalent diseases mainly through sexual transmission, with more than 80% of sexually active people becoming infected with HPV in their lifetime ^3^.

Vaccines are considered one of the most effective public health interventions, saving millions of lives each year. Clinical data has demonstrated that HPV vaccination is most effective in preventing HPV-related cancers among these “unexposed” individuals^4, 5^. World Health Organization (WHO) advocates that the HPV vaccine should be introduced in the national immunization programs (NIP), and recommends girls aged 9-14 years as the primary target population for HPV vaccination^6^. By November 2023, 140 (72%) countries have introduced the HPV vaccine into their NIPs^7^, most of these are high income countries.

The global HPV vaccination coverage of girls that received at least one dose of HPV vaccine has increased to 21% in 2022^7^. Notably, the HPV vaccine coverage in Low- and Middle-income countries is often significantly lower than that in high-income countries^8^. However, despite being economically developed countries, HPV vaccination coverage for girls has not reached the 90% target set by the WHO. The behavior of delaying or refusing vaccines despite vaccine availability, widely known as “vaccine hesitancy”^9–11^, which was listed as one of the top ten health threats by the WHO and became an important reason for the delay in reaching the target value for HPV vaccination coverage^9^.

In 2019, 63% of parents in the United States did not initiate the HPV vaccine for their adolescents aged 13-17^12^. Even by 2022, 23% were still hesitant^13^. Similarly, in 2019, in the UK, 38%^14^ of parents refused or delayed the HPV vaccination for children aged 9-12, and about 32%^15^ of Greek parents have also postponed making a decision regarding the vaccination for their teenagers. In the same year, 59%^16^ of South African public-school parents and 54%^17^ of Chinese parents of teenagers expressed hesitation. Hence, vaccine availability is just one reason on hesitance. Parents play a paramount role as the primary decision makers in regard to vaccinating minors against HPV. The process of making this crucial decision is intricate and influenced by various significant factors, encompassing attitudes, health beliefs, knowledge, sociodemographic, as well as cultural and religious dimensions^18–20^.

China actively responds to the call of WHO to “eliminate cervical cancer” in some regions, although it is not currently one of the NIP countries. Ordos City, Inner Mongolia (an ethnic autonomous region), China, is a prefecture-level city with low health resources, it contains two Districts and seven Banners (similar to counties). In August 2020, the local government of Ordos City has pioneered a program in the Junggar Banner (one of the seven banners): set up 27 vaccination sites to provide free HPV vaccine (Cervarix^®^, GlaxoSmithKline) to all primary and secondary school female students aged 13 years or older (about 8,500). Ordos city has become the first city providing free HPV vaccine in China, which is an important step on China’s path to achieve the goal of eliminating cervical cancer by 2030 as advocated by the WHO.

In 2021, other areas of Ordos City also launched a program to provide free HPV vaccination for girls aged 13-18. In the same year, Xiamen, Jinan, Shenzhen, and Chengdu were designated as the Chinese initial cohort of pilot cities in China to offer free HPV vaccines to schoolgirls. As of August 2023, a total of seven provincial-level regions (including municipalities and autonomous regions) and twenty-seven prefecture-level or district-level regions in China have initiated HPV vaccine vaccination programs that were either free or offered at reduced cost.

Although the vaccinations have generally done well in Ordos City over the past two years, there are still individual schools where there is room for improvement in the current vaccination coverage. Therefore, we conducted a study in Ordos City to explore the potential influencing factors of awareness, knowledge, health beliefs and willingness of parents of girls who were not vaccinated against HPV. We aimed to provide new insights into the development of interventions to increase HPV vaccination rates among adolescents and gather foundational evidence for other newly launched areas in China where HPV vaccine immunization programs were implemented.

## 2 Material and methods

### 2.1 Setting and sampling

We used cluster random sampling to select Banners/Districts of Ordos City. Considering that the HPV vaccination rate of female students at some schools were close to 90%, convenient sampling strategy was adopted to select schools with high parental vaccine hesitation, and the school directors were contacted by the local maternal and child health hospital to obtain approval to distribute questionnaires. In each selected school, classes meeting the inclusion criteria were selected, and the teachers forwarded the questionnaire to the parents. Participants in this study were expected to meet the following criteria: (1) have at least one girl aged 13-18 in the family; (2) the girl had not been vaccinated against the HPV; (3) parents were able to recognize and understand simple words; (4) voluntary participants signed an informed consent form and submitted a completed questionnaire, and those who did not complete the questionnaire were excluded from the study.

The study was approved by the Ethics Committee of Peking Union Medical College [CAMS&PUMC-IEC-2022-077]. Participants were fully informed of the purpose of the study and were invited to participate voluntarily.

Before launching the study, we conducted a pilot trial to determine the feasibility of the study design, evaluating the validity of the indicators and optimizing the study protocol. The pilot study was conducted according to formal research procedures. The valid rate of questionnaires from the pilot trial was as high as 90%, and 451 valid questionnaires were included after 23 repeated and illogical questionnaires were deleted. According to Cronbach’s α test, both the knowledge and the health beliefs section of the questionnaire had good validity (0.7-0.9), and the preliminary analysis of some variables had statistical significance among groups (P<0.05). After conducting the pilot study, we fine-tuned the questionnaire. Therefore, the sample size of the pilot study was not included in the final analysis.

### 2.2 Research questionnaire

By reviewing the literatures^21–24^, a set of electronic questionnaire was developed, which collected information on the following topics: (1) sociodemographics, including age, gender, marital status, daughters’ place of registered permanent residence, number of daughters, own or spouses’ education level and working status, family history of cervical cancer, and their attitude about vaccination to themselves or spouses and daughters; (2) Awareness and knowledge of HPV and HPV vaccine; (3) health beliefs related HPV. The four core dimensions of the Health Belief Model were adopted, namely: perceived susceptibility to HPV and cervical cancer (2 items); perceived severity of HPV infection and cervical cancer (3 items); perceived benefits of vaccine (4 items); perceived barriers to vaccination (3 items). Specifically, perceived susceptibility refers to the belief in the likelihood of a disease occurring. Perceived severity refers to the degree to which the negative effects of the disease are considered severe. Perceived benefit is the belief that a vaccine will reduce the risk or severity of the disease. Perceived barriers refer to any perceived difficulty preventing vaccination. The reliability of these knowledge questions has been validated with a Cronbach’s α = 0.742; the reliability analysis for the Cronbach’s α of the four dimensions of health belief were respectively 0.930, 0.854, 0.844, and 0.736; suggesting a strong reliability.

Regarding the knowledge and health beliefs, all items were rated using “Agree”, “Disagree” or “I don’t know/Unsure” answers that scored 0 (knowledge: wrong answer/ don’t know; health beliefs: disagree or don’t know/ Unsure) or 1 (knowledge: correct answer; health beliefs: agree), except for the dimension of the perceived barrier of health beliefs, which was reversely scored (0 scores: agree or don’t know/Unsure; 1: disagree)^17, 25^. Scores of relevant items were summed to calculate the total score of the knowledge and health belief and then divided by the number of questions to obtain the mean value. Higher total scores and mean scores indicate higher knowledge about HPV and vaccines, and higher beliefs in health behavior.

### 2.3 Data collection

This study was carried out from March to April 2023. Three Banners were included finally: the Dalat Banner, Yijin Horo Banner and Hangjin Banner. Six junior middle or senior high schools from each selected Banner area were selected. Parents of female students aged 13-18 who had not started being vaccinated against HPV were invited to participate in the survey via an electronic questionnaire (www.wjx.com). A maximum of one month to collect the questionnaires was allowed. In total 1818 questionnaires were distributed with 1752 (96.37%) parents providing valid information.

After the questionnaire was completed, logical errors in the questionnaire were checked and corrected, and unqualified surveys were excluded by trained investigators.

### 2.4 Statistical analysis

Descriptive statistics were computed for all parents in different characteristics by number and percentages. Logistic regressions were used to identify factors associated with parents’ awareness and knowledge of the HPV vaccine, health beliefs, and willingness. HPV and HPV vaccine knowledge and health beliefs were assessed using mean scores. The mean knowledge score (KS) and health belief score (HBS) were dichotomized into the categories of ‘‘low (≤ median)” and ‘‘high (> median)”^26, 27^, and sections above mean KS and mean HBS for bivariate and multivariable analysis. Adjusted odds ratio (aOR) and corresponding 95% confidence interval (CI) were presented. Statistical significance was assessed by two-tailed tests with a level of 0.05. Analyses were performed using R software, version 3.5.2.

## 3 Results

### 3.1 Demographics

A total of 1752 girls who had not been vaccinated, and 1547 parents with an average age of 42.25 years (parents with only one girl: 41 years; parents with two or more girls: 43 years) were included in the final analysis after deleting duplicate responses (when the same family answered twice or more) and illogical values (when all the answers were obviously the same). Table1 shows the demographic characteristics and basic information of the subjects. The majority of the parents were female (92.4%), had only one 13-18-year-old daughter (78.6%), were of Han nationality (94.2%), were from urban areas (82.2%), and had an education level higher than junior high school (83.0%). More than half of them were no older than 45 years old (65.7%) and were employed (59.7%).

**Table 1.** Demographic characteristics and basic information of the subjects.

| **characteristics** | **All, N=1547** | **%** |
| --- | --- | --- |
| **Gender of parents** |  |  |
| Male | 118 | 7.6 |
| Female | 1429 | 92.4 |
| **Number of daughters** |  |  |
| ≥ 2 | 331 | 21.4 |
| 1 | 1216 | 78.6 |
| **Age of parents** |  |  |
| <35 | 103 | 6.7 |
| 35-39 | 448 | 29 |
| 40-44 | 465 | 30.1 |
| ≥ 45 | 444 | 28.7 |
| Missing | 87 | 5.6 |
| **Have your daughters registered permanent residence in Ordos?** |  |  |
| No | 427 | 27.6 |
| Yes | 1120 | 72.4 |
| **Marital status** |  |  |
| Never married | 37 | 2.4 |
| Married | 1451 | 93.8 |
| Other | 59 | 3.8 |
| **Ethnicity** |  |  |
| Other | 90 | 5.8 |
| Han | 1457 | 94.2 |
| **Place of permanent residence** |  |  |
| Urban | 1272 | 82.2 |
| Rural | 275 | 17.8 |
| **Educational attainment** |  |  |
| Elementary and below | 263 | 17 |
| Middle school to high school | 921 | 59.5 |
| College (including technical college) and above | 363 | 23.5 |
| **Employment** |  |  |
| Unemployed (including retiree, housewife) | 623 | 40.3 |
| Workers (including self-employed) | 489 | 31.6 |
| Employees (including civil servants, institutions) | 270 | 17.5 |
| Others (including farmers) | 165 | 10.7 |
| **Household income (RMB/month)** |  |  |
| < 6000 | 900 | 58.2 |
| 6000-10000 | 290 | 18.8 |
| >10000 | 140 | 9.1 |
| Missing | 217 | 14 |
| **Family history of cervical cancer** |  |  |
| No | 1488 | 96.2 |
| Yes | 20 | 1.3 |
| Don't know | 39 | 2.5 |
| **Have you/your spouse been vaccinated against HPV?** |  |  |
| No | 1177 | 76.1 |
| Yes | 370 | 23.9 |
| **Are you/your spouse willing to be vaccinated against HPV? (n=1177)** |  |  |
| No | 127 | 8.2 |
| Yes | 827 | 53.5 |
| Unclear | 223 | 14.4 |

### 3.2 Awareness and knowledge

Supplemental Table S1 shows results of the awareness and knowledge survey. Most respondents had heard of HPV (78.1%) and the HPV vaccine (93%). Slightly more parents (15%) had heard of the HPV vaccine than those who had heard of HPV, and the median score for knowledge was 1.98 (SD: 1.81) out of 7. Specifically, among parents who had heard of HPV (n = 1208), around half (44%) knew that the primary mode of transmission of HPV was sexual transmission, and one-third (33.3%) of them perceived that males could also be infected with HPV. Fewer (27.5%) parents agreed that HPV infection could cause other diseases besides cervical cancer. The percentage of parents who knew that HPV infection could occur without any symptoms was no more than one in four (22.6%). Among parents who had heard of the HPV vaccine (n = 1439), around one-fifth (17.3%) agreed with the statement that being infected with HPV means that they must develop cervical cancer. Approximately 35% of parents thought that HPV vaccination was necessary even if HPV infection was present and 34.7% believed that someone being vaccinated against HPV means that they would not develop cervical cancer.

Table 2 shows the multivariable analysis of awareness on HPV vaccine among the participants: the factors associated with increased HPV vaccine awareness were being females (aOR = 3.08, 95% CI: 1.58-6.02), aged 35-39 (aOR = 3.14, 95% CI: 1.17-8.44) or over 45 years old (aOR = 2.60, 95% CI: 1.01-6.69), having a middle school to high school degree (aOR = 2.50, 95% CI: 1.42-4.40), even college or higher degrees (aOR = 4.46, 95% CI: 1.64-12.16), had vaccinated against HPV (aOR = 4.30, 95% CI: 1.66-11.14) or were willing to vaccinate to themselves or their spouses (aOR = 4.64, 95% CI: 2.37-9.10); the factors negatively associated with HPV vaccine awareness were registered permanent residence (girls) (aOR = 0.44, 95% CI: 0.22-0.86) and had farm work (aOR = 0.36, 95% CI: 0.18-0.72). The factors regarding high knowledge level of the HPV vaccine (Table 3), parents who were females (aOR = 1.87, 95% CI: 1.21-2.87), had a college or higher education (aOR = 1.65, 95% CI: 1.05-2.59), and had a monthly household income of 6,000-10,000 RMB (aOR = 1.68, 95% CI: 1.25-2.27) tended to have higher knowledge level; parents who had married (aOR = 0.29, 95% CI: 0.09-0.97) reported lower knowledge scores (Table3).

**Table 2.** Factors associated with HPV vaccine awareness.

| Variables | Had heard of HPV vaccine, N (%), n=1439 | OR (95% CI) | a OR (95% CI) |
| --- | --- | --- | --- |
| **Gender of parents** |  |  |  |
| Male | 99 (83.9) | 1(ref) | 1(ref) |
| Female | 1340 (93.8) | **2.89(1.69-4.94) \*\*** | **3.08(1.58-6.02) \*\*** |
| **Number of daughters** |  |  |  |
| ≥ 2 | 307 (92.7) | 1(ref) | 1(ref) |
| 1 | 1132 (93.1) | 1.05(0.66-1.69) | 1.38(0.75-2.53) |
| **Age of parents** |  |  |  |
| <35 | 93 (90.3) | 1(ref) | 1(ref) |
| 35-39 | 432 (96.4) | **2.90(1.28-6.60) \*** | **3.14(1.17-8.44) \*** |
| 40-44 | 439 (94.4) | 1.82(0.85-3.89) | 2.14(0.83-5.54) |
| ≥ 45 | 399 (89.9) | 0.95(0.46-1.96) | **2.60(1.01-6.69) \*** |
| **Have your daughters registered permanent residence in Ordos?** |  |  |  |
| No | 412 (96.5) | 1(ref) | 1(ref) |
| Yes | 1027 (91.7) | **0.40(0.23-0.70) \*\*** | **0.44(0.22-0.86) \*** |
| **Marital status** |  |  |  |
| Never married | 31 (83.8) | 1(ref) | 1(ref) |
| Married | 1355 (93.4) | **2.73(1.11-6.71) \*** | - |
| Other | 53 (89.8) | 1.71(0.51-5.76) | - |
| **Ethnicity** |  |  |  |
| Other | 84 (93.3) | 1(ref) | 1(ref) |
| Han | 1355 (93) | 0.95(0.41-2.23) | 1.47(0.58-3.72) |
| **Place of permanent residence** |  |  |  |
| Urban | 1203 (94.6) | 1(ref) | 1(ref) |
| Rural | 236 (85.8) | **0.35(0.23-0.53) \*\*** | 1.00(0.54-1.86) |
| **Educational attainment** |  |  |  |
| Elementary and below | 221 (84) | 1(ref) | 1(ref) |
| Middle school to high school | 866 (94) | **2.99(1.95-4.59) \*\*** | **2.50(1.42-4.40) \*\*** |
| College (including technical college) and above | 352 (97) | **6.08(3.07-12.06) \*\*** | **4.46(1.64-12.16) \*** |
| **Employment** |  |  |  |
| Unemployed (including retiree, housewife) | 582 (93.4) | 1(ref) | 1(ref) |
| Workers (including self-employed) | 462 (94.5) | 1.21(0.73-1.99) | 0.85(0.45-1.60) |
| Employees (including civil servants, institution) | 261 (96.7) | 2.04(0.98-4.27) | 0.90(0.34-2.34) |
| Others (including farmers) | 134 (81.2) | **0.31(0.18-0.50) \*\*** | **0.36(0.18-0.72) \*** |
| **Household income (RMB/month)** |  |  |  |
| < 6000 | 833 (92.6) | 1(ref) | 1(ref) |
| 6000-10000 | 274 (94.5) | 1.38(0.79-2.42) | 0.86(0.46-1.62) |
| >10000 | 136 (97.1) | 2.74(0.98-7.62) | 1.73(0.58-5.13) |
| **Family history of cervical cancer** |  |  |  |
| No | 1386 (93.1) | 1(ref) | 1(ref) |
| Yes | 18 (90) | 0.66(0.15-2.89) | 0.27(0.054-1.34) |
| Don't know | 35 (89.7) | 0.64(0.22-1.85) | 0.74(0.23-2.34) |
| **Have you/your spouse been vaccinated against HPV?** |  |  |  |
| No | 1076 (91.4) | 1(ref) | 1(ref) |
| Yes | 363 (98.1) | **4.87(2.24-10.57) \*\*** | **4.30(1.66-11.14) \*\*** |
| **Are you/your spouse willing to be vaccinated against HPV? (n=926)** |  |  |  |
| No | 109 (85.8) | 1(ref) | 1(ref) |
| Yes | 779 (94.2) | **2.68(1.50-4.78) \*\*** | **4.64(2.37-9.10) \*\*** |
| Unclear | 188 (84.3) | 0.89(0.48-1.64) | 0.53(0.26-1.04) |

Bold denotes $p < 0.05$ .
\* $P < 0.05$
\*\* $P < 0.001$

**Table 3.** Factors associated with HPV vaccine knowledge.

| Variables | KS≥2, N(%),<br>n=805 | OR (95% CI) | a OR (95% CI) |
| --- | --- | --- | --- |
| **Gender of parents** |  |  |  |
| Male | 51 (43.2) | 1(ref) | 1(ref) |
| Female | 754 (52.8) | **1.47(1.01-2.14) \*** | **1.87(1.21-2.87) \*\*** |
| **Number of daughters** |  |  |  |
| ≥ 2 | 163 (49.2) | 1(ref) | 1(ref) |
| 1 | 642 (52.8) | 1.15(0.90-1.47) | 0.96(0.71-1.29) |
| **Age of parents** |  |  |  |
| <35 | 55 (53.4) | 1(ref) | 1(ref) |
| 35-39 | 225 (50.2) | 0.88(0.57-1.35) | 0.96(0.56-1.64) |
| 40-44 | 260 (55.9) | 1.11(0.72-1.70) | 1.12(0.65-1.93) |
| ≥ 45 | 226 (50.9) | 0.91(0.59-1.39) | 1.22(0.71-2.11) |
| **Have your daughters registered permanent residence in Ordos?** |  |  |  |
| No | 217 (50.8) | 1(ref) | 1(ref) |
| Yes | 588 (52.5) | 1.07(0.86-1.34) | 0.96(0.74-1.24) |
| **Marital status** |  |  |  |
| Never married | 23 (62.2) | 1(ref) | 1(ref) |
| Married | 751 (51.8) | 0.65(0.33-1.28) | **0.29(0.09-0.97) \*** |
| Other | 31 (52.5) | 0.67(0.29-1.56) | 0.37(0.10-1.40) |
| **Ethnicity** |  |  |  |
| Other | 51 (56.7) | 1(ref) | 1(ref) |
| Han | 754 (51.8) | 0.82(0.53-1.26) | 1.12(0.70-1.84) |
| **Place of permanent residence** |  |  |  |
| Urban | 682 (53.6) | 1(ref) | 1(ref) |
| Rural | 123 (44.7) | **0.70(0.54-0.91) \*\*** | 0.95(0.68-1.33) |
| **Educational attainment** |  |  |  |
| Elementary and below | 122 (46.4) | 1(ref) | 1(ref) |
| Middle school to high school | 427 (46.4) | 1.00(0.76-1.32) | 0.83(0.60-1.16) |
| College (including technical college) and above | 256 (70.5) | **2.77(1.99-3.85) \*\*** | **1.65(1.05-2.59) \*** |
| **Employment** |  |  |  |
| Unemployed (including retiree, housewife) | 280 (44.9) | 1(ref) | 1(ref) |
| Workers (including self-employed) | 264 (54) | **1.44(1.13-1.82) \*\*** | 1.17(0.88-1.55) |
| Employees (including civil servants, institutions) | 181 (67) | **2.49(1.85-3.36) \*\*** | 1.30(0.87-1.93) |
| Others (including farmers) | 80 (48.5) | 1.15(0.82-1.63) | 0.88(0.58-1.34) |
| **Household income (RMB/month)** |  |  |  |
| < 6000 | 427 (47.4) | 1(ref) | 1(ref) |
| 6000-10000 | 187 (64.5) | **2.01(1.53-2.64) \*\*** | **1.68(1.25-2.27) \*\*** |
| >10000 | 89 (63.6) | **1.93(1.34-2.79) \*\*** | 1.40(0.93-2.10) |
| **Family history of cervical cancer** |  |  |  |
| No | 771 (51.8) | 1(ref) | 1(ref) |
| Yes | 13 (65) | 1.73(0.69-4.35) | 2.80(0.87-8.99) |
| Don't know | 21 (53.8) | 1.09(0.57-2.05) | 0.89(0.44-1.81) |
| **Have you/your spouse been vaccinated against HPV?** |  |  |  |
| No | 577 (49) | 1(ref) | 1(ref) |
| Yes | 228 (61.6) | **1.67(1.32-2.12) \*\*** | 1.23(0.92-1.63) |
| **Are you/your spouse willing to be vaccinated against HPV? (n=926)** |  |  |  |
| No | 61 (48) | 1(ref) | 1(ref) |
| Yes | 436 (52.7) | 1.21(0.83-1.75) | 1.22(0.79-1.90) |
| Unclear | 80 (35.9) | **0.61(0.39-0.94) \*** | 0.74(0.44-1.25) |

Bold denotes $p < 0.05$ .
\* $P < 0.05$
\*\* $P < 0.001$

### 3.3 Health beliefs

With regard to the health beliefs, the median score was 6.19 (SD: 2.94) of 12 (Table 4). Among the subjects, 58.7% had a high perceived susceptibility, 19.5% had a high perceived severity, 55.4% had a high perceived barrier, and 68.1% had a high perceived benefit. Specifically, less than one-fifth of parents thought that their daughters were at risk for HPV infection (17.5%) or cervical cancer (17%). Most parents perceived that being infected with HPV was scary (70%:) and that it could have a significant impact on their daughter’s life (76.2%:), and cervical cancer could devastate their families (69.9%). Over half of the parents expressed the HPV vaccine was important (72%) or necessary (67.9%) for their daughters. Many parents believed that being vaccinated could reduce the risk of HPV infection (59.1%) or cervical cancer (79.9%). Less than half of the parents trusted the safety (39.7%), effectiveness (33.1%), and absence of side effects (17.3%) of the HPV vaccine.

**Table 4.** Factors associated with health beliefs.

| Variables | Health beliefs Score > 6 ,<br>N(%), n=816 | OR (95% CI) | a OR (95% CI) |
| --- | --- | --- | --- |
| **Gender of parents** |  |  |  |
| Male | 64 (54.2) | 1(ref) | 1(ref) |
| Female | 752 (52.6) | 0.94(0.64-1.37) | 0.96(0.63-1.46) |
| **Number of daughters** |  |  |  |
| ≥ 2 | 173 (52.3) | 1(ref) | 1(ref) |
| 1 | 643 (52.9) | 1.03(0.80-1.31) | 0.86(0.64-1.15) |
| **Age of parents** |  |  |  |
| <35 | 54 (52.4) | 1(ref) | 1(ref) |
| 35-39 | 235 (52.5) | 1.00(0.65-1.54) | 0.83(0.49-1.42) |
| 40-44 | 253 (54.4) | 1.08(0.71-1.66) | 0.88(0.51-1.50) |
| ≥ 45 | 235 (52.9) | 1.02(0.66-1.57) | 0.96(0.56-1.65) |
| **Have your daughters registered permanent residence in Ordos?** |  |  |  |
| No | 254 (59.5) | 1(ref) | 1(ref) |
| Yes | 562 (50.2) | **0.69(0.55-0.86) \*\*** | **0.61(0.47-0.80) \*\*** |
| **Marital status** |  |  |  |
| Never married | 19 (51.4) | 1(ref) | 1(ref) |
| Married | 767 (52.9) | 1.06(0.55-2.04) | 1.17(0.41-3.34) |
| Other | 30 (50.8) | 0.98(0.43-2.23) | 1.26(0.38-4.14) |
| **Ethnicity** |  |  |  |
| Other | 45 (50) | 1(ref) | 1(ref) |
| Han | 771 (52.9) | 1.12(0.73-1.72) | 1.10(0.69-1.76) |
| **Place of permanent residence** |  |  |  |
| Urban | 681 (53.5) | 1(ref) | 1(ref) |
| Rural | 135 (49.1) | 0.84(0.65-1.09) | 0.95(0.69-1.32) |
| **Educational attainment** |  |  |  |
| Elementary and below | 124 (47.1) | 1(ref) | 1(ref) |
| Middle school to high school | 468 (50.8) | 1.15(0.88-1.52) | 0.96(0.69-1.34) |
| College (including technical college) and above | 224 (61.7) | **1.81(1.31-2.49) \*\*** | 1.31(0.84-2.03) |
| **Employment** |  |  |  |
| Unemployed (including retiree, housewife) | 301 (48.3) | 1(ref) | 1(ref) |
| Workers (including self-employed) | 257 (52.6) | 1.19(0.94-1.50) | 1.03(0.77-1.37) |
| Employees (including civil servants, institutions) | 165 (61.1) | **1.68(1.26-2.25) \*\*** | 1.22(0.83-1.80) |
| Others (including farmers) | 93 (56.4) | 1.38(0.98-1.95) | 1.36(0.90-2.07) |
| **Household income (RMB/month)** |  |  |  |
| < 6000 | 451 (50.1) | 1(ref) | 1(ref) |
| 6000-10000 | 167 (57.6) | **1.35(1.04-1.77) \*** | 1.23(0.92-1.64) |
| >10000 | 91 (65) | **1.85(1.28-2.68) \*\*** | **1.67(1.11-2.50) \*** |
| **Family history of cervical cancer** |  |  |  |
| No | 780 (52.4) | 1(ref) | 1(ref) |
| Yes | 12 (60) | 1.36(0.55-3.35) | 1.98(0.67-5.82) |
| Don't know | 24 (61.5) | 1.45(0.76-2.79) | 1.46(0.72-2.98) |
| **Have you/your spouse been vaccinated against HPV?** |  |  |  |
| No | 590 (50.1) | 1(ref) | 1(ref) |
| Yes | 226 (61.1) | **1.56(1.23-1.98) \*\*** | 1.20(0.91-1.59) |
| **Are you/your spouse willing to be vaccinated against HPV? (n=926)** |  |  |  |
| No | 57 (44.9) | 1(ref) | 1(ref) |
| Yes | 467 (56.5) | **1.59(1.09-2.32) \*** | **1.87(1.20-2.91) \*\*** |
| Unclear | 66 (29.6) | **0.52(0.33-0.81) \*\*** | **0.52(0.30-0.89) \*** |

Bold denotes $p < 0.05$ .
\* $P < 0.05$
\*\* $P < 0.001$

Multivariate logistic regression (Table 4) showed that parents’ health beliefs increased with monthly household income, especially for those with a monthly income of >10,000 RMB (aOR = 1.67, 95% CI: 1.11-2.50). Those who were willing to vaccinate themselves or their spouse against HPV had higher health beliefs (aOR = 1.87, 95% CI: 1.20-2.91). In contrast, parents whose daughters with permanent residence in Ordos City had lower health beliefs (aOR = 0.61, 95% CI: 0.47-0.80).

### 3.4 Willingness

Willingness towards vaccination was presented in Table 5. The results showed a high level of parents’ willingness to have their daughters receive the HPV vaccination (85.5%). The multivariate regression analysis indicated that parents whose daughters had permanent residence in Ordos City were less willing to accept vaccination (aOR = 0.54, 95% CI: 0.35-0.82), and parents with a monthly household income of 6,000-10,000 RMB (aOR = 1.72, 95% CI: 1.06-2.81) were more inclined to accept vaccines for their daughters. Additionally, parental willingness to vaccinate themselves or their spouses was positively correlated with their willingness to vaccinate their daughters (aOR = 6.09, 95% CI: 3.48-10.65).

**Table 5.** Factors associated with willingness to get HPV vaccination for daughters.

| Variables | Willinging (%), n=1323 | OR (95% CI) | a OR (95% CI) |
| --- | --- | --- | --- |
| **Gender of parents** |  |  |  |
| Male | 103 (87.3) | 1 (Ref) | 1 (Ref) |
| Female | 1220 (85.4) | 0.85(0.49-1.49) | 0.97(0.52-1.81) |
| **Number of daughters** |  |  |  |
| ≥ 2 | 280 (84.6) | 1 (Ref) | 1 (Ref) |
| 1 | 1043 (85.8) | 1.10(0.78-1.54) | 0.99(0.65-1.53) |
| **Age of parents** |  |  |  |
| <35 | 86 (83.5) | 1 (Ref) | 1 (Ref) |
| 35-39 | 394 (87.9) | 1.44(0.80-2.61) | 1.65(0.78-3.47) |
| 40-44 | 394 (84.7) | 1.10(0.62-1.96) | 1.07(0.52-2.23) |
| ≥ 45 | 380 (85.6) | 1.17(0.66-2.10) | 1.46(0.69-3.07) |
| **Have your daughters registered permanent residence in Ordos?** |  |  |  |
| No | 381 (89.2) | 1 (Ref) | 1 (Ref) |
| Yes | 942 (84.1) | **0.64(0.45-0.90) \*** | **0.54(0.35-0.82) \*** |
| **Marital status** |  |  |  |
| Never married | 29 (78.4) | 1 (Ref) | 1 (Ref) |
| Married | 1243 (85.7) | 1.65(0.74-3.66) | 0.70(0.14-3.50) |
| Other | 51 (86.4) | 1.76(0.60-5.18) | 0.82(0.13-5.04) |
| **Ethnicity** |  |  |  |
| Other | 75 (83.3) | 1 (Ref) | 1 (Ref) |
| Han | 1248 (85.7) | 1.20(0.67-2.12) | 1.12(0.58-2.16) |
| **Place of permanent residence** |  |  |  |
| Urban | 1097 (86.2) | 1 (Ref) | 1 (Ref) |
| Rural | 226 (82.2) | 0.74 (0.52-1.04) | 0.86(0.54-1.36) |
| **Educational attainment** |  |  |  |
| Elementary and below | 214 (81.4) | 1 (Ref) | 1 (Ref) |
| Middle school to high school | 794 (86.2) | 1.43(1.00-2.06) | 1.32(0.84-2.08) |
| College (including technical college) and above | 315 (86.8) | 1.50(0.97-2.32) | 1.12(0.60-2.09) |
| **Employment** |  |  |  |
| Unemployed (including retiree, housewife) | 526 (84.4) | 1 (Ref) | 1 (Ref) |
| Workers (including self-employed) | 419 (85.7) | 1.10(0.79-1.54) | 0.97(0.64-1.47) |
| Employees (including civil servants, institutions) | 241 (89.3) | 1.53(0.99-2.38) | 1.34(0.74-2.44) |
| Others (including farmers) | 137 (83) | 0.90(0.57-1.43) | 1.13(0.62-2.04) |
| **Household income (RMB/month)** |  |  |  |
| < 6000 | 765 (85) | 1 (Ref) | 1 (Ref) |
| 6000-10000 | 264 (91) | **1.79(1.15-2.79) \*** | **1.72(1.06-2.81) \*** |
| >10000 | 120 (85.7) | 1.06(0.64-1.76) | 0.97(0.56-1.70) |
| **Family history of cervical cancer** |  |  |  |
| No | 1271 (85.4) | 1 (Ref) | 1 (Ref) |
| Yes | 19 (95) | 3.46(0.39-30.90) | 2.02(0.26-15.68) |
| Don't know | 33 (84.6) | 1.07(0.44-2.57) | 0.77(0.31-1.94) |
| **Have you/your spouse been vaccinated against HPV?** |  |  |  |
| No | 990 (84.1) | 1 (Ref) | 1 (Ref) |
| Yes | 333 (90) | **1.70(1.17-2.47) \*\*** | 1.53(0.98-2.34) |
| **Are you/your spouse willing to be vaccinated against HPV? (n=926)** |  |  |  |
| No | 86 (67.7) | 1 (Ref) | 1 (Ref) |
| Yes | 761 (92) | **5.50(3.51-8.61) \*\*** | **6.09(3.48-10.65) \*\*** |
| Unclear | 143 (64.1) | 0.85(0.54-1.35) | 0.60(0.34-1.04) |

Bold denotes $p < 0.05$ .
\* $P < 0.05$
\*\* $P < 0.001$

### 3.5 Information source

The results showed that the main sources of information regarding the HPV vaccine were traditional media (52.3%) and family/ friends/colleagues (48.7%) (Supplemental Figure S1). Hospitals (48.6%) were the most trusted institutions by parents, followed by schools (36.1%) (Supplemental Figure S2). The top concerns for parents regarding the acceptance of HPV vaccines were “safety” (38.8%), and 29% (28.8%) of respondents expressed concerns about “all aspects of the vaccines” (Supplemental Figure S3).

## 4 Discussion

To the best of our knowledge, this study was the first in China to assess the influencing factors of knowledge and health beliefs related to HPV, as well as the willingness of parents to vaccinate their daughters after the introduction of the free HPV vaccine in Ordos City. In total, 1547 parents whose daughters had not been vaccinated were analyzed finally, 78% of them had heard of HPV, 93% had heard of the HPV vaccine, and 85.5% were willing to vaccinate their daughters. Factors that potentially influence parents’ knowledge level were found to be gender, married status, education, and income. Registered permanent residence (girls), income, and vaccination status (female parents) were significantly associated with parental health beliefs and willingness to vaccinate daughters.

The awareness level of HPV (78%) and HPV vaccines (93%) of participants in this study was higher compared to that of previous Chinese (61.02% and 51.11% in 2019^17^) and some NIP countries’ (such as the United Kingdom^14^: both 55% in 2019 and Poland^28^: 74.2% and 61.4% in 2022) reported results. Different levels of awareness also could be observed in different parts of the same country or city. For instance, a conspicuous discrepancy in awareness regarding HPV and its vaccine surfaced between urban and rural areas in Zhejiang, China^29^ (HPV: 60.8% vs 12.3%; HPV vaccine: 85.0% vs 13.8%); the extent of HPV vaccine awareness displayed inconsistency between the southern and northwestern regions of Ethiopia (85%^30^ vs 73%^25^). This suggests that there is still a lot of work to be done in promoting the HPV vaccine. Our participants mainly received HPV vaccine information from traditional media, as well as from their family, friends, and co-workers. However, they considered hospitals to be the most trusted and expected sources of information, although they were rarely accessible. Indeed, HPV vaccine information can influence an individual’s health behavior and serve as a prompt for action; thus, it is important to consider the advice of healthcare providers when making decisions about the HPV vaccine^18^.

Despite the growing awareness of HPV and the HPV vaccines^31^, insufficient knowledge and understanding regarding HPV were prevalent among parents worldwide^32–38^. Our subjects had an overall low level of knowledge and demonstrated a lack of understanding even on basic questions. For instance, nearly a half (44%) of the parents were unaware that sexual transmission is the most common route of transmission of HPV, and about two thirds (77%) of parents did not know that the men also could be infected with HPV. And even the healthcare providers, there was room for improvement on knowledge and understanding of HPV and its vaccines^39, 40^. Knowledge is an important factor influencing attitude and behavior^41^. Therefore, the urgent need to take effective health education measures for different groups and provide personalized health care services for them.

Our findings were consistent with previous research in demonstrating that gender, education, income, and marital status were associated with parents’ knowledge of HPV and its vaccines. Specifically, we observed that male participants had lower knowledge related to HPV compared to females^25, 42^. This may be more common in countries where the HPV vaccine is only provided for females; males in these countries could be neglected in daily vaccine promotion and education. Based on this situation, future health education on HPV should consider including males and preparing for the eventual introduction of the HPV vaccine for them. Moreover, higher family income and education level were positively correlated with parents’ knowledge level^27, 43–45^. Individuals with higher income were more likely to focus on their own health development^46^ and have better access to healthcare and resources. Similarly, individuals with a higher education level have greater comprehension skills, making it easier for them to access reliable public health information^27, 45^. Furthermore, a significant association between being married status and insufficient knowledge was found. Our participants were mostly married and generally younger, which means that they were basically married early. People who marry early usually have low education level, leaving them with limited opportunities for education and self-development, thus, more attention and support should be devoted to their health education^47, 48^.

Ordos parents’ willingness to vaccinate their daughters was as high as 85.5%, surpassing other regions in China such as Zhejiang (35.4%)^49^, as well as the provinces of Sichuan, Henan, and Heilongjiang provinces; (46.4%)^17^. It was also comparable to some areas in other countries, like Buenos Aires City in Argentina (88.5%)^50^ and Wroclaw in Poland (86.8%)^51^. As other studies have shown^52, 53^, higher income has been found to have a positive influence, whereas higher educational level in our study has not been found to have the same impact as previous research. Thus, further investigation was necessary. Interestingly, in our study, high health beliefs and vaccination willingness share influential factors. This suggests that there could be a correlation between health beliefs and willingness to be vaccinated. Indeed, wealthier people often have more access to health resources and more likely to form positive health beliefs and attitudes. Notably, the percentages of perceived susceptibility, perceived severity, and perceived benefit among our participants were not optimistic. Enhancing people’s knowledge of these aspects is an important task we are facing. In other words, there is an urgent need for health education in these areas to raise public awareness of diseases or risks. Although about half of the parents do not have a high level of perceived barriers, which is beneficial for our health intervention work, it is still an issue that needs to be addressed.

Our findings also revealed some novel findings about health beliefs and willingness. Parents’ health beliefs and their willingness to vaccinate themselves was highly positively correlated with their willingness to vaccinate their girls, indicating that parental attitudes could influence the healthy behavior of the two female generations in the family. This emphasize that the importance of family or mother-daughter joint forms of interventions. Moreover, compared to parents of girls with non-local household, parents of girls with local household had lower health beliefs and were more likely to be hesitant about the HPV vaccine. This further underscores the necessity for local health education efforts. Perhaps as an area with ethnic characteristics, Ordos City’s unique values and customs also cannot be ignored^54, 55^. Therefore, targeted education and publicity efforts are needed to raise the health awareness and attitudes of the target population.

In our results, barriers to vaccination were included concerns about vaccine safety, efficacy, side effects, and uncertainty about how to schedule an appointment, with safety being the most important concern for parents. More and more reports of adverse events after immunization have made people start to panic about vaccination^56^, leading to people becoming more cautious about vaccination^57–60^. Nevertheless, data indicates the HPV vaccine in global use is safe currently ^61^. Thus, disseminating correct and reliable information about HPV vaccines is important.

### Strength and limitations

Our study’s principal strength was that it was the first survey conducted in the first area where free HPV vaccination was already available in China. We explore parental awareness, knowledge, health beliefs, and willingness regarding HPV and HPV vaccines in Ordos City, can provide foundational evidence for another new HPV vaccine immunization program implementation area, and generate ideas for effective interventions. It could provide some data support and guidance significance for Chinese NIP work in the future. It also has certain limitations. Mainly the health belief modeling tool for HPV vaccination intention lacked standardization, the web-based questionnaire lacked flexibility; and the sample size was insufficient on subgroups, such as gender and ethnicity to explore the influence of different characteristics.

## 5 Conclusions

In conclusion, factors that were found to potentially influence knowledge included gender, married status, education, and income. Registered permanent residence (girls), income, and vaccination status (female parents) were significantly associated with health beliefs and willingness. There may be a correlation between vaccine intention and health beliefs. Parental attitudes could influence the healthy behavior of the entire family. It is worth highlighting the importance of improving the accuracy and correctness of health education content and the professionalism of education personnel to address issues of low health knowledge and belief.

## 6 Statements and Declarations

### 6.1 Competing interests

The authors have no relevant financial or non-financial interests to disclose.

### 6.2 Author contribution

Study concepts and Study design: JY and Y-LQ; Data acquisition: S-ST, JY, Y-X, L-LZ, J-X; Quality control data and algorithms: S-ST and JY; Data analysis, interpretation and statistical analysis: S-ST, JY and Y-FL; Manuscript preparation: S-ST, JY, Y-FL, S-MW and H-JY; Manuscript editing: S-ST, JY and Y-FL; Manuscript review: JY, X-ML and Y-LQ; All authors read and approved the final manuscript.

### 6.3 Data availability

The data presented in this study are fully included in the manuscript and supplementary materials, with no additional data required for replication of the findings. Raw data generated during the study are available from the corresponding author upon reasonable request.

### 6.4 Ethics approval

The study was approved by the Ethics Committee of Peking Union Medical College [CAMS&PUMC-IEC-2022-077].

### 6.5 Funding

This research was supported by grants from CAMS Innovation Fund for Medical Sciences (CAMS2021-12M-1-004).

## Notes

### Competing Interest Statement

The authors have declared no competing interest.

### Funding Statement

This study was funded by CAMS Innovation Fund for Medical Sciences (CAMS2021-12M-1-004)

### Author Declarations

Ethics committee of Chinese Academy of Medical Sciences and Peking Union Medical College gave ethical approval for this work

