## Supplementary tables and figures for "Factors on Knowledge, Health Beliefs, and Willingness toward Human Papillomavirus Vaccine among Chinese Parents, a cross-sectional study in Ordos City, China"

| **Supplementary material e Table 1 Awareness, knowledge, and health beliefs of HPV and HPV vaccine.** | | |
| --- | --- | --- |
| **Variables** | **N** | **%** |
| **Awareness** |  |  |
| Whether heard of HPV |  |  |
| No | 339 | 21.9 |
| Yes | 1208 | 78.1 |
| Whether heard of the HPV vaccine |  |  |
| No | 108 | 7 |
| Yes | 1439 | 93 |
| **Knowledge (mean KS: 1.98 ± 1.81)** |  |  |
| HPV is mainly through sexual transmission (n = 1208). |  |  |
| Disagree | 168 | 13.9 |
| Agree | 532 | 44 |
| Unsure | 508 | 42.1 |
| Only women could be infected with HPV (n = 1208). |  |  |
| Disagree | 402 | 33.3 |
| Agree | 302 | 25 |
| I don't know | 504 | 41.7 |
| HPV infection may occur without any symptoms (n = 1208). |  |  |
| Disagree | 352 | 29.1 |
| Agree | 273 | 22.6 |
| I don't know | 583 | 48.3 |
| HPV infection does not only lead to cervical cancer (n = 1208). |  |  |
| Disagree | 274 | 22.7 |
| Agree | 332 | 27.5 |
| I don't know | 602 | 49.8 |
| Being infected with HPV means that you must develop cervical cancer. (n = 1208) |  |  |
| Disagree | 243 | 20.1 |
| Agree | 209 | 17.3 |
| I don't know | 756 | 62.6 |
| If you are infected with HPV, there is no need to get the HPV vaccine (n = 1439). |  |  |
| Disagree | 503 | 35 |
| Agree | 231 | 16.1 |
| I don't know | 705 | 49 |
| Being vaccinated against HPV means that you won't develop cervical cancer. (n = 1439). |  |  |
| Disagree | 500 | 34.7 |
| Agree | 102 | 7.1 |
| I don't know | 837 | 58.2 |
| **Health beliefs (n = 1547) (mean HBS: 6.19 ± 2.94)** |  |  |
| **Perceived susceptibility (n = 908, 58.7%) (mean: 2.16 ± 1.15)** |  |  |
| My children are at risk for HPV infection. |  |  |
| Disagree | 546 | 35.3 |
| Agree# | 270 | 17.5 |
| Unsure | 731 | 47.3 |
| My children are at risk of cervical cancer. |  |  |
| Disagree | 578 | 37.4 |
| Agree# | 263 | 17 |
| Unsure | 706 | 45.6 |
| **Perceived severity (n = 301, 19.5%) (mean: 0.35 ± 0.73)** |  |  |
| Infected HPV could have a significant impact on my children’s life. |  |  |
| Disagree | 84 | 5.4 |
| Agree# | 1179 | 76.2 |
| Unsure | 284 | 18.4 |
| The thought of my children getting HPV is scary. |  |  |
| Disagree | 157 | 10.1 |
| Agree# | 1083 | 70 |
| Unsure | 307 | 19.8 |
| Cervical cancer will devastate my family and my children. |  |  |
| Disagree | 147 | 9.5 |
| Agree# | 1081 | 69.9 |
| Unsure | 319 | 20.6 |
| **Perceived barriers (n = 857, 55.4%) (mean: 0.90 ± 1.13)** |  |  |
| I doubt the safety of the vaccine. |  |  |
| Disagree# | 614 | 39.7 |
| Agree | 207 | 13.4 |
| Unsure | 726 | 46.9 |
| I doubt the effectiveness of the vaccine. |  |  |
| Disagree# | 512 | 33.1 |
| Agree | 230 | 14.9 |
| Unsure | 805 | 52 |
| I doubt the vaccines have side effects. |  |  |
| Disagree# | 267 | 17.3 |
| Agree | 211 | 13.6 |
| Unsure | 1069 | 69.1 |
| **Perceived benefits (n = 1053, 68.1%) (mean: 2.79 ± 1.40)** |  |  |
| HPV vaccines are very important. |  |  |
| Disagree | 80 | 5.2 |
| Agree# | 1114 | 72 |
| Unsure | 353 | 22.8 |
| HPV vaccines are necessary. |  |  |
| Disagree | 99 | 6.4 |
| Agree# | 1051 | 67.9 |
| Unsure | 397 | 25.7 |
| The HPV vaccine can protect my children from HPV. |  |  |
| Disagree | 99 | 6.4 |
| Agree# | 915 | 59.1 |
| Unsure | 533 | 34.5 |
| The HPV vaccine can reduce my children's risk of cervical cancer. |  |  |
| Disagree | 53 | 3.4 |
| Agree# | 1233 | 79.7 |
| Unsure | 261 | 16.9 |

To better complete this survey, the "HPV vaccine" appeared in the form of "cervical cancer vaccine" in our questionnaire.

“Strongly disagree” and “disagree” were combined to be “disagree”; “strongly agree” and “agree” were combined to be “agree.”

#: The answer is correct, or it favors the happen of vaccination behavior.

|  | Percent (%) |
| --- | --- |
| Traditional media | 52.3 |
| Family/ friends/ colleague | 48.7 |
| Others | 43.2 |
| Social media | 32.6 |
| healthcare workers | 25.4 |

**Supplementary material e Figure 1 What are your sources of information on the HPV vaccine?**

|  | Percent (%) |
| --- | --- |
| Hospitals | 48.6 |
| School | 36.1 |
| All | 24.6 |
| CDC | 22.1 |
| Communities | 10.4 |
| None | 3.7 |

**Supplementary material e Figure 2 Which organization do you trust more for vaccination?**

CDC: Centers for Disease Control and Prevention

|  | Percent (%) |
| --- | --- |
| Safety | 38.8 |
| All | 28.8 |
| Others | 16.3 |
| Effectiveness | 14 |
| None | 2.2 |

**Supplementary material e Figure 3 What are your concerns about the HPV vaccine?**
